# SARS-CoV-2 infections in migrants and the role of household overcrowding: A causal mediation analysis of Virus Watch data

**DOI:** 10.1101/2022.12.21.22283794

**Authors:** Yamina Boukari, Sarah Beale, Vincent Grigori Nguyen, Wing Lam Erica Fong, Rachel Burns, Alexei Yavlinsky, Susan Hoskins, Kate Marie Lewis, Cyril Geismar, Annalan M D Navaratnam, Isobel Braithwaite, Thomas E Byrne, Youssof Oskrochi, Sam Tweed, Jana Kovar, Parth Patel, Andrew C Hayward, Robert W Aldridge

## Abstract

**Background:** Migrants are over-represented in severe acute respiratory syndrome coronavirus-2 (SARS-CoV-2) infections globally; however, evidence is limited for migrants in England and Wales. Household overcrowding is a risk factor for SARS-CoV-2 infection, with migrants more likely to live in overcrowded households than UK-born individuals. We aimed to estimate the total effect of migration status on SARS-CoV-2 infection and to what extent household overcrowding mediated this effect.

**Methods:** We included a sub-cohort of individuals from the Virus Watch prospective cohort study during the second SARS-CoV-2 wave (1st September 2020–30th April 2021) who were aged ≥18 years, self-reported the number of rooms in their household and had no evidence of SARS-CoV-2 infection pre-September 2020. We estimated total, indirect and direct effects using Buis’ logistic decomposition regression controlling for age, sex, ethnicity, clinical vulnerability, occupation, income and whether they lived with children.

**Results:** In total, 23,478 individuals were included. 9.07% (187/2,062) of migrants had evidence of infection during the study period versus 6.27% (1,342/21,416) of UK-born individuals. Migrants had 22% higher odds of infection during the second wave (total effect; OR:1.22, 95%CI:1.01–1.47). Household overcrowding accounted for approximately 32% of these increased odds (indirect effect, OR:1.07, 95%CI:1.03–1.12; proportion accounted for: indirect effect[7]/total effect[22]=0.32).

**Conclusion:** Migrants had higher odds of SARS-CoV-2 infection during the second wave compared with UK-born individuals and household overcrowding explained 32% of these increased odds. Policy interventions to reduce household overcrowding for migrants are needed as part of efforts to tackle health inequalities during the pandemic and beyond.

**Key messages:** *What is already known on this topic:*

- Migrants in England and Wales may be at greater risk of exposure to SARS-CoV-2 due to unique risk factors, including over-representation in front-line jobs, an increased likelihood of living in multigenerational households and difficulties in accessing primary care. Research shows that migrants in high-income countries have been disproportionally infected with SARS-CoV-2. It is likely that, due to their pre-existing vulnerabilities, this is similarly the case for migrants in England and Wales; however, quantitative evidence addressing this is lacking.

*What this study adds:*

- We investigated the effect of being a migrant on SARS-CoV-2 infection during the second wave of the pandemic in a cohort in England and Wales. We also determined the proportion of the effect mediated by household overcrowding after controlling for age, sex, ethnicity, clinical vulnerability, occupation, income and the presence of children in the household. Migrants had 22% higher odds of being infected with SARS-CoV-2 than their UK-born counterparts, and household overcrowding accounted for approximately 32% of these increased odds.

*How this study might affect research, practice or policy:*

- Our findings highlight the role of household overcrowding in the disproportionate impact of SARS-CoV-2 infections on migrants. They also demonstrate the urgent need for policy interventions that improve housing conditions as part of efforts to reduce health inequalities. Further research investigating other causes of migrants’ over-representation in infection cases is also needed to inform further targeted policy interventions.

## Background

Globally, the UK has the fifth largest migrant (non UK-born) population comprising approximately 9.57 million people in 2020^1, 2^. Migrants in the UK may be at greater risk of SARS-CoV-2 infection due to pre-existing vulnerabilities such as their over-representation in front-line jobs (e.g. in healthcare, hospitality, retail and delivery sectors), increased use of public transport and increased likelihood of living in multi-generational households^3, 4^. Barriers to accessing primary care are well documented for migrants^5-9^ and may negatively impact vaccine uptake, thus potentially putting migrants at greater risk of infection and severe outcomes from the combination of greater exposure and under-vaccination^10^. Migrants in high-income countries have been over-represented in SARS-CoV-2 infections, hospitalisations and deaths^11-13^. UK-focused quantitative evidence is limited but does suggest inequalities. In England, a study showed a greater increase in all-cause mortality for migrants versus non-migrants from 21 March to 8 May 2020 when compared with previous years’ deaths^14^.

The built environment is a wider determinant of health^15^. Household overcrowding is associated with a risk to physical and mental health, and is a potential marker of social deprivation^16^. Growing UK-focused evidence links household overcrowding to SARS-CoV-2 infection and other COVID-19-related outcomes^17, 18^. In England and Wales, individuals who participated in the Virus Watch study and lived in overcrowded households had higher odds of testing positive for SARS-CoV-2 infection via polymerase chain reaction (PCR) and antibody tests than individuals living in underoccupied households^17^. Similar findings were reported from the COVIDENCE UK study^18^ and studies that utilised related measures such as household size when controlling for various demographic, social, behavioural and comorbidity characteristics^19-22^ or area-level housing indicators^23^. Household size also played a role in differences in COVID-19 outcomes for South Asian groups after adjusting for sociodemographic and clinical factors^24, 25^.

Household overcrowding is particularly relevant to migrants. In London, 13–16% of migrant households were overcrowded compared to just 4% of UK-born households between 2016 to 2018^26^. Outside of London, the overcrowding rates were lower with 2% of UK-born households being overcrowded compared to 5–8% of migrant households. Despite the lack of UK-focused studies, in Europe and the US, household overcrowding is a reported risk factor for SARS-CoV-2 exposure in migrants^27-29^, thus highlighting the need for investigation in a UK-based sample. We aimed to examine the odds of SARS-CoV-2 infection for migrants versus UK-born individuals during the second COVID-19 wave, and whether household overcrowding mediated the effect of migration status on SARS-CoV-2 infection.

## Method

### Study setting

We used data from Virus Watch, a prospective community cohort study of COVID-19 in England and Wales from 1st June 2020^30^. Virus Watch included 58,628 individuals as of 28th July 2022. Our analysis was restricted to the second wave, from 1st September 2020–30th April 2021^31^, as migrants were likely to have high exposure risk early in the pandemic and because testing was not widespread during the first wave. Households were recruited from 24th June 2020 to March 2022 and asked to complete a post-enrolment baseline survey containing demographic, medical history, financial and occupation questions. Individuals received a weekly illness survey via email to collect information on self-reported acute symptoms, vaccination status and PCR or lateral flow test results. Households also received a monthly survey of demographic, health-related, environmental and behavioural/psychosocial questions. Within the larger study, a sub-cohort of adults (the laboratory cohort) received monthly antibody testing.

Virus Watch cohort data were linked to the Second-Generation Surveillance System (SGSS) containing laboratory SARS-CoV-2 test results from swabs taken during hospitalisation (Pillar 1) and community testing (Pillar 2)^32, 33^. The linkage period was March 2020–August 2021 for Pillar 1, and June 2020–November 2021 for Pillar 2. Linkage was conducted by NHS Digital using the name, date of birth, address and NHS number variables, which were sent in March 2021.

### Participants

Participants were aged ≥18 years and reported the number of rooms in their household in the February 2021 survey. Participants with evidence of SARS-CoV-2 infection before the start of the second wave (September 2020) were excluded as first infection, rather than reinfection, was our focus.

### Exposure and outcome

Country of birth was the exposure, defined as migrant (i.e. a non-UK country of birth reported at enrolment) or UK-born (a UK country of birth). The outcome was evidence of first SARS-CoV-2 infection during the analysis period defined as either a PCR or lateral flow test self-reported during a weekly survey, a positive test for SARS-CoV-2 anti-nucleocapsid antibodies, a positive test for anti-spike antibodies or a positive PCR test identified via the SGSS.

### Potential mediator: Household overcrowding status

In the February 2021 survey, participating households were asked how many rooms were available for their exclusive use (excluding bathrooms, toilets, halls, landings and cupboards). Persons per room (PPR) was calculated by dividing the total number of people in the household (including children) by the number of available rooms, excluding bathrooms or kitchens. For households reporting ≥2 rooms, one room was subtracted from the total, assuming that one of the rooms was a kitchen. For households reporting one room, it was assumed that this room is likely to be a bedroom/studio. Households with PPRs less than one were defined as ‘under-occupied’, equal to one as ‘balanced’ and greater than one as ‘overcrowded’^17^. The PPR approach is a validated overcrowding measure that has fair agreement with other measures^34^ and has previously been used to determine overcrowding status in Virus Watch^17^.

#### Confounders

Potential confounders were identified using a directed acyclic graph (DAG; Supplementary Figure 1)^35^. To provide minimally adjusted unbiased estimates of the total, indirect and direct effects, we controlled for baseline age, sex at birth, ethnicity (White British, White Irish, White Other, Mixed, South Asian, Other Asian, Black, Other and Unknown), clinical vulnerability (“Not clinically vulnerable”, “Clinically vulnerable”, “Clinically extremely vulnerable” and “Missing” based on self-reported conditions or medications indicating vulnerability using government criteria, adapted for the clinical variables collected at baseline^36, 37^), baseline total household income (£0–9,999, £10,000– 24,999, £25,000–49,000, £50,000–74,999, £75,000–99,999 and £100,000+) and occupation (see Supplementary Box 1 for details).

**Figure 1:**
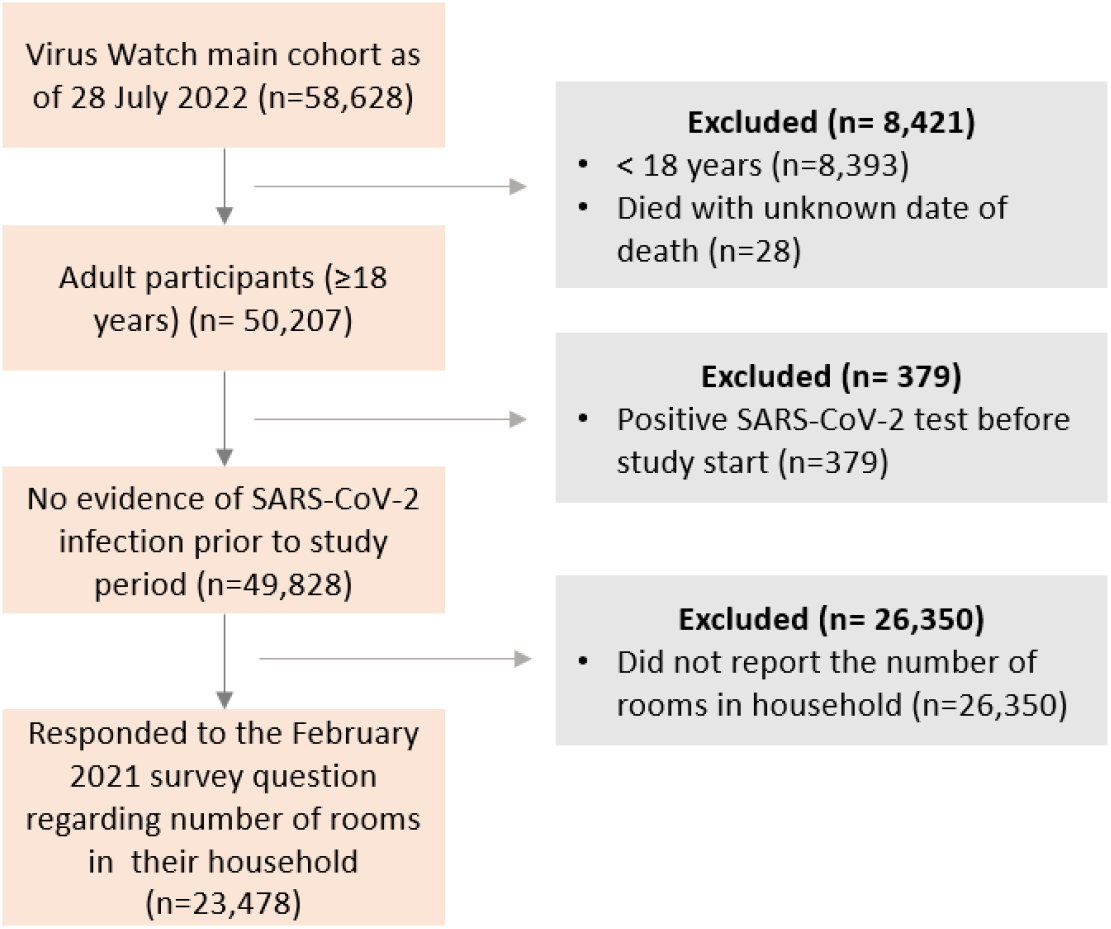
Flow diagram of participant eligibility.

#### Other demographic and clinical characteristics

Households were assigned a geographic region (nine English regions and Wales)^38^ and a local area-level Index of Multiple Deprivation (IMD) quintile (where one represents the most deprived and five the least) by linking their postcode to the May-2020 ONS Postcode Lookup^39^.

### Statistical analysis

We found no evidence of co-linearity between ethnicity and migrant status (Supplementary Box 2). We used Buis’ logistic decomposition regression with bootstrapped standard errors to estimate the total and direct effects of migration status on infection, and the indirect effect mediated through household overcrowding^40^ (see Supplementary Box 2 for further methodological information). The percentage of the total effect mediated by household overcrowding was estimated using the indirect effect odds ratio (OR) as the numerator and the total effect OR as the denominator. As the model derives the total effect coefficient by summing the direct and indirect effects on the log scale, the percentage is an approximation only (i.e. indirect and direct effect ORs do not sum to give exactly the total effect).

### Sensitivity analyses

As eligibility was not dependent on households responding to all weekly surveys throughout the analysis period, we conducted a sensitivity analysis including only participants who had either linked data or, for those without linked data, self-reported follow-up for every full week of the analysis period (although this may also bias towards households who were healthy enough to respond each week). Positive SARS-CoV-2 anti-nucleocapsid antibody or anti-spike antibody tests during the study period may indicate evidence of older SARS-CoV-2 infection prior to the second wave or post-vaccination seroconversions. Therefore, we carried out a sensitivity analysis using only swab-confirmed infections We also conducted a sensitivity analysis where household overcrowding was represented by the continuous PPR variable.

Individuals with missing country-of-birth responses were classified as UK-born; however, in a sensitivity analysis, we excluded these individuals. There were no missing age data and 363 participants (1.5%) had missing data for sex, 478 (2.0%) for ethnicity, 1,405 (6.0%) for clinical vulnerability, 4,120 (17.5%) for household income and 3,400 (14.5%) for occupation. All missing values were included in the model under a ‘Missing’ category. We conducted sensitivity analyses to assess the effect of missing data using a complete case analysis and multivariate imputation by chained equations using the *mice* package with 5 datasets and 50 iterations per dataset (see Supplementary Box 2 for the included predictor variables).

Based on *a priori* assumptions, we modelled ethnicity as a confounder of the effect of migration status on infection. However, ethnicity is complex and can encompass country of birth^41^, which creates overlap with our migrant exposure. To explore this, we conducted a sensitivity analysis without adjusting for ethnicity.

### Tools

R version 4.1.2 was used for data cleaning and multiple imputation. Mediation analysis was carried out using the *ldecomp* command in Stata version 17.0.

## Results

Of 58,628 individuals in the Virus Watch cohort on 28th July 2022, 23,478 (40.0%) individuals met our inclusion criteria (Figure 1).

### Demographic characteristics

Migrants were generally younger (median age: 53 versus 63 years in migrants and non-migrants, respectively) with a higher percentage identifying as female (59.1% female migrants versus 54.1% female non-migrants). Migrants identified predominantly with a minority ethnic group (75.6%) versus White British (23.5%) and were less likely to have missing ethnicity data than UK-born individuals. Over 40% of migrants were situated in London compared with only 9.5% of UK-born individuals, whilst over 40% of the UK-born individuals were in the East or South East of England. Migrants generally lived in more deprived areas compared with the UK-born group, but more migrants lived in households with higher total incomes. Missing income data were more common for UK-born individuals. The percentages of clinically and extremely clinically vulnerable individuals in each group were similar, with more missing data in the UK-born group. Higher percentages of migrants worked in all occupations versus UK-born individuals, apart from in outdoor trade- and transport and machines-related occupations. More UK-born individuals were not in employment versus migrants and missing occupation data were more common in the UK-born versus migrant group.

In the migrant group, the median number of rooms per household was 5 compared to 6 in the UK-born group (Table 1). 10.9% of migrants lived in overcrowded housing compared with 2.0% of the UK-born group. Migrants were less likely to live in under-occupied housing than UK-born individuals (71.4% versus 91.8%, respectively).

**Table 1:** Cohort demographic and household characteristics.

| Characteristic | Overall<br>N = 23,478 | UK-born<br>N = 21,416 | Migrant<br>N = 2,062 |
| --- | --- | --- | --- |
| **Age** |  |  |  |
| Mean (SD) | 59 (15) | 59 (15) | 53 (16) |
| Median (IQR) | 62 (19) | 63 (19) | 53 (26) |
| <30 | 1,379 (5.9%) | 1,241 (5.8%) | 138 (6.7%) |
| 30-39 | 1,718 (7.3%) | 1,341 (6.3%) | 377 (18.3%) |
| 40-49 | 2,537 (10.8%) | 2,163 (10.1%) | 374 (18.1%) |
| 50-59 | 4,397 (18.7%) | 4,022 (18.8%) | 375 (18.2%) |
| 60+ | 13,447 (57.3%) | 12,649 (59.1%) | 798 (38.7%) |
| **Sex** |  |  |  |
| Female | 12,810 (54.6%) | 11,592 (54.1%) | 1,218 (59.1%) |
| Male | 10,276 (43.8%) | Omitted to avoid disclosure | Omitted to avoid disclosure |
| Other or unknown | 392 (1.7%) | Omitted to avoid disclosure | Omitted to avoid disclosure |
| **Ethnicity** |  |  |  |
| White British | 20,456 (87.1%) | 19,972 (93.3%) | 484 (23.5%) |
| Minority ethnic* | 2,544 (10.8%) | 986 (4.6%) | 1,558 (75.6%) |
| Missing | 478 (2.0%) | 458 (2.1%) | 20 (1.0%) |
| **Geographical region** |  |  |  |
| East Midlands | 2,057 (8.8%) | 1,971 (9.2%) | 86 (4.2%) |
| East of England | 5,208 (22.2%) | 4,889 (22.8%) | 319 (15.5%) |
| London | 2,933 (12.5%) | 2,044 (9.5%) | 889 (43.1%) |
| North East | 1,153 (4.9%) | 1,105 (5.2%) | 48 (2.3%) |
| North West | 2,500 (10.6%) | 2,409 (11.2%) | 91 (4.4%) |
| South East | 4,524 (19.3%) | 4,147 (19.4%) | 377 (18.3%) |
| South West | 1,752 (7.5%) | 1,641 (7.7%) | 111 (5.4%) |
| Wales | 540 (2.3%) | 521 (2.4%) | 19 (0.9%) |
| West Midlands | 1,276 (5.4%) | 1,218 (5.7%) | 58 (2.8%) |
| Yorkshire and The Humber | 1,153 (4.9%) | 1,106 (5.2%) | 47 (2.3%) |
| Missing | 382 (1.6%) | 365 (1.7%) | 17 (0.8%) |
| **IMD quintile** |  |  |  |
| 1 (most deprived) | 1,909 (8.1%) | 1,679 (7.8%) | 230 (11.2%) |
| 2 | 3,384 (14.4%) | 2,948 (13.8%) | 436 (21.1%) |
| 3 | 4,770 (20.3%) | 4,297 (20.1%) | 473 (22.9%) |
| 4 | 6,124 (26.1%) | 5,675 (26.5%) | 449 (21.8%) |
| 5 (least deprived) | 6,909 (29.4%) | 6,452 (30.1%) | 457 (22.2%) |
| Missing | 382 (1.6%) | 365 (1.7%) | 17 (0.8%) |
| **Total household income** |  |  |  |
| £0-9,999 | 806 (3.4%) | 712 (3.3%) | 94 (4.6%) |
| £10,000-24,999 | 4,453 (19.0%) | 4,158 (19.4%) | 295 (14.3%) |
| £25,000-49,999 | 7,080 (30.2%) | 6,570 (30.7%) | 510 (24.7%) |
| £50,000-£74,999 | 3,568 (15.2%) | 3,173 (14.8%) | 395 (19.2%) |
| £75,000-£99,999 | 1,726 (7.4%) | 1,504 (7.0%) | 222 (10.8%) |
| £100,000+ | 1,725 (7.3%) | 1,410 (6.6%) | 315 (15.3%) |
| Missing | 4,120 (17.5%) | 3,889 (18.2%) | 231 (11.2%) |
| **Number of rooms in household** |  |  |  |
| Mean (SD) | 6 (2) | 6 (2) | 5 (2) |
| Median (IQR) | 6 (2) | 6 (2) | 5 (3) |
| **Number of householders** |  |  |  |
| 1 | 4,640 (19.8%) | 4,230 (19.8%) | 410 (19.9%) |
| 2 | 13,571 (57.8%) | 12,538 (58.5%) | 1,033 (50.1%) |
| 3 | 2,647 (11.3%) | 2,345 (10.9%) | 302 (14.6%) |
| 4 | 1,983 (8.4%) | 1,738 (8.1%) | 245 (11.9%) |
| 5 | 540 (2.3%) | 478 (2.2%) | 62 (3.0%) |
| 6 | 97 (0.4%) | 87 (0.4%) | 10 (0.5%) |
| **Household overcrowding status** |  |  |  |
| Under occupied | 21,134 (90.0%) | 19,662 (91.8%) | 1,472 (71.4%) |
| Balanced | 1,686 (7.2%) | 1,320 (6.2%) | 366 (17.7%) |
| Overcrowded | 658 (2.8%) | 434 (2.0%) | 224 (10.9%) |
| **Persons per room** |  |  |  |
| Mean (SD) | 0.49 (0.33) | 0.47 (0.29) | 0.71 (0.53) |
| Median (IQR) | 0.40 (0.31) | 0.40 (0.29) | 0.50 (0.67) |
| **Living with any children** |  |  |  |
| No | 20,246 (86.2%) | 18,647 (87.1%) | 1,599 (77.5%) |
| Yes | 3,232 (13.8%) | 2,769 (12.9%) | 463 (22.5%) |
| **Occupation** |  |  |  |
| Administrative and secretarial | 1,378 (5.9%) | 1,238 (5.8%) | 140 (6.8%) |
| Healthcare | 735 (3.1%) | 629 (2.9%) | 106 (5.1%) |
| Indoor trades, process and plant | 660 (2.8%) | 587 (2.7%) | 73 (3.5%) |
| Leisure and personal Service | 484 (2.1%) | 412 (1.9%) | 72 (3.5%) |
| Managers, directors and senior officials | 855 (3.6%) | 760 (3.5%) | 95 (4.6%) |
| Other professional and associate | 3,345 (14.2%) | 2,829 (13.2%) | 516 (25.0%) |
| Outdoor trades | 254 (1.1%) | 232 (1.1%) | 22 (1.1%) |
| Sales and customer service | 521 (2.2%) | 474 (2.2%) | 47 (2.3%) |
| Social care and community protective services | 543 (2.3%) | 485 (2.3%) | 58 (2.8%) |
| Teaching, education and childcare | 1,150 (4.9%) | 1,019 (4.8%) | 131 (6.4%) |
| Transport and mobile machine | 229 (1.0%) | 210 (1.0%) | 19 (0.9%) |
| Not in employment<sup>†</sup> | 9,924 (42.3%) | 9,247 (43.2%) | 677 (32.8%) |
| Missing | 3,400 (14.5%) | 3,294 (15.4%) | 106 (5.1%) |
| **Clinical vulnerability status** |  |  |  |
| Not clinically vulnerable | 12,964 (55.2%) | 11,715 (54.7%) | 1,249 (60.6%) |
| Clinically vulnerable | 6,395 (27.2%) | 5,846 (27.3%) | 549 (26.6%) |
| Clinically extremely vulnerable | 2,714 (11.6%) | 2,470 (11.5%) | 244 (11.8%) |
| Missing | 1,405 (6.0%) | 1,385 (6.5%) | 20 (1.0%) |

| **Characteristic** | **Overall**<br>**N = 23,478** | **UK-born**<br>**N = 21,416** | **Migrant**<br>**N = 2,062** |
| --- | --- | --- | --- |

\*All minority ethnic groups were combined to avoid statistical disclosure. <sup>†</sup>Represents individuals who are retired, students, looking after the household/family and not looking for work, permanently sick or disabled, or unemployed.

### Evidence of SARS-CoV-2 infection

From 1st September 2020 to 30th April 2021, 1,529/23,478 individuals had evidence of SARS-CoV-2 infection (Table 2). In both groups, evidence of infection was identified via a swab test in >50% of positive cases, with a slightly higher percentage of swab tests in migrants versus UK-born individuals (55.1% versus 53.1%, respectively).

**Table 2:** Source of positive test results for all participants with first evidence of SARS-CoV-2 infection within the analysis period (1^St^ September 2020 to 30^th^ April 2021)

| Source of positive test result | Overall<br>*N* = 1,529 | UK-born<br>*n*=1,342 (87.8%) | Migrant<br>*n*=187 (12.2%) |
| --- | --- | --- | --- |
| Antibody tests* | 713 (46.6%) | 629 (46.9%) | 84 (44.9%) |
| Swab tests (PCR or lateral flow) | 816 (53.4%) | 713 (53.1%) | 103 (55.1%) |

\*Tests for SARS-CoV-2 anti-nucleocapsid antibodies and anti-spike antibodies.

In the migrant group, 9.07% (187/2,062) of individuals had evidence of infection compared with 6.27% (1,342/21,416) of UK-born individuals (Table 3). In both groups, the percentage of participants with evidence of infection was highest in individuals living in overcrowded housing (migrant: 15.2% [34/224]; UK-born: 9.9% [43/434]) compared with individuals living in under-occupied housing (migrant: 6.9% [102/1,472]; UK-born: 6.0% [1,185/19,662]).

**Table 3:** Percentage of participants with evidence of SARS-CoV-2 infection in the second wave (1 September 2020 to 30 April 2021)

| Characteristic | UK-born |  |  | Migrant |  |  |
| --- | --- | --- | --- | --- | --- | --- |
|  | Overall,<br>N=21,416<sup>1</sup> | Evidence of SARS-CoV-2 infection |  | Overall,<br>N = 2,062 | Evidence of SARS-CoV-2 infection |  |
|  |  | No<br>n=20,074 (93.7%) | Yes<br>n=1,342 (6.27%) |  | No<br>n=1,875 (90.9%) | Yes<br>n=187 (9.07%) |
| Age |  |  |  |  |  |  |
| <30 | 1,241 | 1,125 (90.7%) | 116 (9.3%) | 138 | 123 (89.1%) | 15 (10.9%) |
| 30-39 | 1,341 | 1,235 (92.1%) | 106 (7.9%) | 377 | 332 (88.1%) | 45 (11.9%) |
| 40-49 | 2,163 | 1,969 (91.0%) | 194 (9.0%) | 374 | 322 (86.1%) | 52 (13.9%) |
| 50-59 | 4,022 | 3,691 (91.8%) | 331 (8.2%) | 375 | 343 (91.5%) | 32 (8.5%) |
| 60+ | 12,649 | 12,054 (95.3%) | 595 (4.7%) | 798 | 755 (94.6%) | 43 (5.4%) |
| Sex |  |  |  |  |  |  |
| Male | 9,435 | 8,877 (94.1%) | 558 (5.9%) | 841 | 770 (91.6%) | 71 (8.4%) |
| Female | 11,592 | 10,823 (93.4%) | 769 (6.6%) | 1,218 | 1,102 (90.5%) | 116 (9.5%) |
| Ethnicity |  |  |  |  |  |  |
| White British | 19,972 | 18,723 (93.7%) | 1,249 (6.3%) | 484 | 451 (93.2%) | 33 (6.8%) |
|  | Overall,<br>N=21,416<sup>1</sup> | Evidence of SARS-CoV-2 infection |  | Overall,<br>N = 2,062 | Evidence of SARS-CoV-2 infection |  |
|  |  | No<br>n=20,074 (93.7%) | Yes<br>n=1,342 (6.27%) |  | No<br>n=1,875 (90.9%) | Yes<br>n=187 (9.07%) |
| Minority ethnic | 986 | 910 (92.3%) | 76 (7.7%) | 1,558 | 1,404 (90.1%) | 154 (9.9%) |
| Missing | 458 | 441 (96.3%) | 17 (3.7%) | 20 | 20 (100.0%) | 0 (0.0%) |
| **Geographical region<sup>1</sup>** |  |  |  |  |  |  |
| East of England | 4,889 | 4,611 (94.3%) | 278 (5.7%) | 319 | 297 (93.1%) | 22 (6.9%) |
| London | 2,044 | 1,830 (89.5%) | 214 (10.5%) | 889 | 769 (86.5%) | 120 (13.5%) |
| South East | 4,147 | 3,927 (94.7%) | 220 (5.3%) | 377 | 358 (95.0%) | 19 (5.0%) |
| **IMD quintile** |  |  |  |  |  |  |
| 1 (most deprived) | 1,679 | 1,526 (90.9%) | 153 (9.1%) | 230 | 211 (91.7%) | 19 (8.3%) |
| 2 | 2,948 | 2,730 (92.6%) | 218 (7.4%) | 436 | 375 (86.0%) | 61 (14.0%) |
| 3 | 4,297 | 4,054 (94.3%) | 243 (5.7%) | 473 | 435 (92.0%) | 38 (8.0%) |
| 4 | 5,675 | 5,334 (94.0%) | 341 (6.0%) | 449 | 417 (92.9%) | 32 (7.1%) |
| 5 (least deprived) | 6,452 | 6,075 (94.2%) | 377 (5.8%) | 457 | 420 (91.9%) | 37 (8.1%) |
|  | Overall,<br>N=21,416<sup>1</sup> | Evidence of SARS-CoV-2 infection |  | Overall,<br>N = 2,062 | Evidence of SARS-CoV-2 infection |  |
|  |  | No<br>n=20,074 (93.7%) | Yes<br>n=1,342 (6.27%) |  | No<br>n=1,875 (90.9%) | Yes<br>n=187 (9.07%) |
| Missing | 365 | 355 (97.3%) | 10 (2.7%) | 17 | 17 (100.0%) | 0 (0.0%) |
| **Total household income** |  |  |  |  |  |  |
| £0-9,999 | 712 | 666 (93.5%) | 46 (6.5%) | 94 | 81 (86.2%) | 13 (13.8%) |
| £10,000-24,999 | 4,158 | 3,915 (94.2%) | 243 (5.8%) | 295 | 271 (91.9%) | 24 (8.1%) |
| £25,000-49,999 | 6,570 | 6,140 (93.5%) | 430 (6.5%) | 510 | 474 (92.9%) | 36 (7.1%) |
| £50,000-£74,999 | 3,173 | 2,914 (91.8%) | 259 (8.2%) | 395 | 360 (91.1%) | 35 (8.9%) |
| £75,000-£99,999 | 1,504 | 1,391 (92.5%) | 113 (7.5%) | 222 | 198 (89.2%) | 24 (10.8%) |
| £100,000+ | 1,410 | 1,308 (92.8%) | 102 (7.2%) | 315 | 277 (87.9%) | 38 (12.1%) |
| Missing | 3,889 | 3,740 (96.2%) | 149 (3.8%) | 231 | 214 (92.6%) | 17 (7.4%) |
| **Household overcrowding status** |  |  |  |  |  |  |
| Under-occupied | 19,662 | 18,477 (94.0%) | 1,185 (6.0%) | 1,472 | 1,370 (93.1%) | 102 (6.9%) |
| Balanced | 1,320 | 1,206 (91.4%) | 114 (8.6%) | 366 | 315 (86.1%) | 51 (13.9%) |
|  | Overall,<br>N=21,416<sup>1</sup> | Evidence of SARS-CoV-2 infection |  | Overall,<br>N = 2,062 | Evidence of SARS-CoV-2 infection |  |
|  |  | No<br>n=20,074 (93.7%) | Yes<br>n=1,342 (6.27%) |  | No<br>n=1,875 (90.9%) | Yes<br>n=187 (9.07%) |
| Overcrowded | 434 | 391 (90.1%) | 43 (9.9%) | 224 | 190 (84.8%) | 34 (15.2%) |
| **Lives with any children** |  |  |  |  |  |  |
| No | 18,647 | 17,530 (94.0%) | 1,117 (6.0%) | 1,599 | 1,472 (92.1%) | 127 (7.9%) |
| Yes | 2,769 | 2,542 (91.8%) | 227 (8.2%) | 463 | 403 (87.0%) | 60 (13.0%) |
| **Clinical vulnerability status** |  |  |  |  |  |  |
| Not clinically vulnerable | 11,715 | 10,946 (93.4%) | 769 (6.6%) | 1,249 | 1,123 (89.9%) | 126 (10.1%) |
| Clinically vulnerable | 5,846 | 5,457 (93.3%) | 389 (6.7%) | 549 | 513 (93.4%) | 36 (6.6%) |
| Clinically extremely vulnerable | 2,470 | 2,345 (94.9%) | 125 (5.1%) | 244 | 219 (89.8%) | 25 (10.2%) |
| Missing | 1,385 | 1,326 (95.7%) | 59 (4.3%) | 20 | 20 (100.0%) | 0 (0.0%) |

<sup>1</sup>Only the geographical areas with the highest proportion of individuals are included to avoid statistical disclosure.

### Causal mediation analysis

Migrants had 22% higher odds of SARS-CoV-2 infection during the second wave versus UK-born individuals (total effect), determined using logistic decomposition regression adjusted for age, sex, ethnicity, clinical vulnerability, baseline total household income and occupation (OR:1.22, 95%CI:1.01–1.47, p=0.041; Table 4). An OR of 1.07 (95%CI: 1.03–1.12, p=0.002) for the indirect effect indicates that household overcrowding partially mediated the relationship between migration status and SARS-CoV-2 infection, accounting for approximately 32% of the total effect (7/22 multiplied by 100). A positive, but statistically insignificant direct effect of migration status on SARS-CoV-2 infection remained after accounting for the indirect effect of household overcrowding status (OR: 1.13, 95%CI: 0.94–1.37, p=0.198).

**Table 4:** ORs for total, indirect and direct effects of migration status on SARS-CoV-2 infection.

| Effect | OR | 95% CI | P value |
| --- | --- | --- | --- |
| Total | 1.22 | 1.01–1.47 | 0.041 |
| Indirect | 1.07 | 1.03–1.12 | 0.002 |
| Direct | 1.13 | 0.94–1.37 | 0.198 |

### Sensitivity analyses

Consistent indirect effect sizes were observed in all the sensitivity analyses, with varying statistical significance (Table 5). Total effects and direct effects were also generally consistent across sensitivity analyses, with the exception of the complete case analysis and after excluding individuals with missing country of birth, likely due to power issues with the reduced sample size (Table 5).

**Table 5:** Total, indirect and direct effects from sensitivity analyses.

| Sensitivity analysis | N | Effect | OR | 95% CI | P value |
| --- | --- | --- | --- | --- | --- |
| Outcome-related |  |  |  |  |  |
| Restricted denominator | 22,496 | Total | 1.17 | 0.97–1.41 | 0.098 |
|  |  | Indirect | 1.07 | 1.02–1.12 | 0.003 |
|  |  | Direct | 1.09 | 0.90–1.32 | 0.368 |
| Positive swab tests only | 23,479 | Total | 1.20 | 0.92–1.56 | 0.174 |
|  |  | Indirect | 1.12 | 1.07–1.18 | <0.001 |
|  |  | Direct | 1.07 | 0.83–1.37 | 0.609 |
| Exposure-related |  |  |  |  |  |
| Excluding individuals with unknown country of birth | 20,748 | Total | 1.04 | 0.85–1.28 | 0.687 |
|  |  | Indirect | 1.06 | 1.02–1.10 | 0.002 |
|  |  | Direct | 0.98 | 0.81–1.20 | 0.872 |
| Mediator-related |  |  |  |  |  |
| Use of PPR as mediator | 23,478 | Total | 1.23 | 1.03–1.46 | 0.023 |
|  |  | Indirect | 1.07 | 1.03–1.11 | <0.001 |
|  |  | Direct | 1.14 | 0.96–1.36 | 0.132 |
| Confounder-related |  |  |  |  |  |
| Multiple imputation of missing confounder data | 23,478 | Total | 1.21 | 0.99–1.48 | 0.067 |
|  |  | Indirect | 1.07 | 1.03–1.11 | 0.001 |
|  |  | Direct | 1.13 | 0.92–1.38 | 0.238 |
| Complete case analysis | 18,466 | Total | 1.07 | 0.86– 1.33 | 0.550 |
|  |  | Indirect | 1.07 | 1.01–1.12 | 0.011 |
|  |  | Direct | 1.00 | 0.80–1.26 | 0.982 |
| Without adjusting for ethnicity | 23,478 | Total | 1.24 | 1.05–1.47 | 0.011 |
|  |  | Indirect | 1.08 | 1.03–1.12 | <0.001 |
|  |  | Direct | 1.16 | 0.97–1.37 | 0.095 |

## Discussion

We present findings indicating that migrants had 22% higher odds of being infected with SARS-CoV-2 during the second wave of the pandemic compared to UK-born individuals after controlling for baseline demographic, socioeconomic and clinical confounders. This increased odds of infection aligns with findings from other high-income countries showing migrants’ over-representation in SARS-CoV-2 infections^11^ and is likely due to amplified pre-pandemic inequalities. These findings, alongside evidence showing migrants’ low COVID-19 vaccine uptake^10, 42^, highlight the need to carefully consider delivery and prioritisation of booster COVID-19 vaccines.

We found a significant positive indirect effect, with household overcrowding explaining approximately 32% of the increased odds of SARS-CoV-2 infection in migrants compared with UK-born individuals, which is consistent with Norwegian and American studies linking household overcrowding and SARS-CoV-2 infections^27, 28^. A direct effect was found after accounting for mediation by household overcrowding, consistent with complementary mediation whereby the investigated mediator has a significant causal role alongside other unmeasured variables^43^.

A strength of this analysis is the focus on a causal mechanism underlying migrants’ increased SARS-CoV-2 infection odds in a substantial group of over 2,000 migrants, allowing for specific policy recommendations to help reduce health inequalities. The use of a DAG-informed model facilitated comprehensive adjustment for confounders and interpretation of the mediated effects. Other strengths are the inclusion of self-reported and linked data on SARS-CoV-2 infection, which reduced reliance on participant recall, and the use of multiple sensitivity analyses.

A limitation is that migrants in Virus Watch are not representative of England’s migrant population. Lead householders who spoke English and had internet access were eligible, whereas evidence suggests that vulnerable, marginalised migrants have limited access to technology and experience English difficulties^44^. Additionally, only households of ≤6 people were eligible, which may induce selection bias given that migrants are more likely to live in larger, multi-generational households^3^. We also focused on infections that occurred during the second wave and excluded individuals who were infected in the first wave, which could potentially induce bias given the risk factors for higher exposure faced by migrants versus UK-born individuals early in the pandemic^12^. Consequently, it is likely that we underestimated the effect of migration status on infection and the household overcrowding-mediated indirect effect.

Individuals whose country of birth was unknown (n=2,730) were classified as UK-born. Whilst this could introduce misclassification bias whereby true migrants are classified as being UK-born, the impact is likely small and would underestimate the effect of migration status on infection. Additionally, results from the sensitivity analysis excluding individuals with a missing country of birth were generally consistent with the main analysis.

We did not adjust for vaccination status at baseline as no individuals had been vaccinated at the start of the analysis period (roll-out began in England on 8th December 2021). However, by 25th April 2021, 91.5% (22,644,679) of individuals aged ≥45 years had received at least one dose^45^. Evidence suggests that vaccination status is a separate mediator of SARS-CoV-2 infection in migrants, with differential uptake of COVID-19 vaccines across migrant and UK-born groups and under-immunisation in migrants in Europe for both COVID-19 and routine vaccinations^42^. Consequently, adjustment for vaccination status should not influence the indirect effect through household overcrowding.

Other limitations are the exclusion of children (<18 years), a group which requires further research. Additionally, Virus Watch enrolled more individuals aged over 60 years or of White British ethnicity versus England’s and Wales’s general population, and included more higher income households. These biases likely contribute to an underestimation of the effect of migration status on infection.

The disproportionate effect of household overcrowding on individuals in the migrant group compared with UK-born individuals builds upon previous Virus Watch results showing household overcrowding as a risk factor for SARS-CoV-2 infection^17^. Household overcrowding has become more common over recent years, particularly in the private and social rental sectors^46, 47^. Migrants are more likely to privately rent their homes and have lower rates of homeownership than non-migrants^26^, which may explain the differential impact on this group. Findings from the two analyses demonstrate the health implications of existing housing inequalities. They highlight the importance of addressing overcrowding as part of a public health strategy to reduce health inequalities, and to ensuring the UK’s preparedness for any subsequent waves or future pandemics. Future research is required to examine other potential mediators of the total effect of migration status on infection to better inform targeted policy interventions across the wider determinants of health.

Efforts to address overcrowding are complex and require engagement with multiple stakeholders, including both in government and the private sector. Short-term efforts to prevent spread include providing hotel accommodation for infected individuals^48^ and ensuring that existing advice on preventing spread within the household (e.g. masking, adequate ventilation etc.)^49^ is accessible to migrant communities. In the medium-term, the statutory overcrowding standard that was introduced in 1935 should be revised as the threshold for breaching it is high, with relatively few households found to be statutorily overcrowded, which limits the ability of local authorities to act^50^. In the long-term, the current housing stock should be reformed. A policy focus in recent years has been on boosting the supply of privately owned homes and increasing home ownership, with a substantial drop in the availability of social housing^47^. Increasing the social housing supply could provide more regulated, secure and affordable housing, whilst enabling swifter improvements in response to policy interventions compared with private housing.

To conclude, we show that migrants were over-represented in SARS-CoV-2 infections early in the pandemic, with household overcrowding playing a significant role in driving over-representation. Our findings highlight the implications of inadequate housing on health and underscore the importance of policy interventions for tackling household overcrowding. As we continue to live with COVID-19, it will be important to address these inequalities in health outcomes and housing to ensure than we build back fairer^51^ and that we are better prepared for future waves and pandemics.

## Data Availability

Data from the Virus Watch cohort are available on the ONS Secure Research Service. The data are available under restricted access as they contain sensitive health data. Access can be obtained via the ONS Secure Research Service.

## Contributors

Conceptualization: YB, RWA. Methodology: YB, SB, VGN, WLEF, KML, AY, SH. Formal Analysis: YB. Data curation: VGN, SB, WLEF, SH. Writing—original draft preparation: YB. Writing—review and editing: YB, SB, VGN, WLEF, RB, AY, SH, KML, CG, IB, TEB, YO, ST, JK, PP, AMDN, ACH, RWA. Visualization: YB. Supervision: RWA, SB, VGN. Project administration: YB. Funding acquisition: RWA, ACH.

## Ethics

Virus Watch was approved by the Hampstead NHS Health Research Authority Ethics Committee (20/HRA/2320) and conformed to the ethical standards set out in the Declaration of Helsinki. Participants provided written informed consent for the collection and use of data prior to study participation.

