## Supplementary Information for "SARS-CoV-2 infections in migrants and the role of household overcrowding: A causal mediation analysis of Virus Watch data"

### Supplementary Box 1: Occupation categories

- Administrative and secretarial
- Healthcare
- Indoor trades, process and plant
- Leisure and personal service
- Managers, directors and senior officials
- Other professional and associate
- Outdoor trades
- Sales and customer service
- Social care and community protective services
- Teaching, education and childcare
- Transport and mobile machine
- Not in employment

### Supplementary Box 2: Causal mediation analysis and multicollinearity checks

An observational study can generally be conceptualised as a conditionally randomised experiment if four key identifiability conditions are met^1^. We outline these conditions and how they relate to our study in Table 1 below.

**Table 1: Identifiability conditions**

| Condition | Met | Rationale |
| --- | --- | --- |
| **No interference** (i.e. an individual’s potential outcome does not depend on another individual’s exposure) | Yes | Another individual being a migrant does not affect whether a UK-born individual gets infected.  However, a study in the US found higher rates of COVID-19 mortality in urban areas with more immigrants who had traditional/multigenerational family structures. Being a migrant is associated with an increased odds of infection and as SARS-CoV-2 is infectious, living in an area with more migrants could mean increased risk of infection for UK-born too. |
| **Consistency** (i.e. the effect of an exposure is the same for all individuals who receive that exposure) | Yes | Migration status is based on whether someone reports a UK or non-UK country of birth. It is not a composite variable. |
| **Positivity** (i.e. all individuals have a probability of greater than 0 of being assigned all values of the exposure variable in every stratum defined by covariates) | Yes | Theoretically possible for all. |
| **Conditional exchangeability** (i.e. all individuals with and without exposure are comparable in every respect apart from the exposure after conditioning on the confounders) | Yes | We controlled for age, sex, ethnicity, clinical vulnerability, household income, occupation and living with children status. |

Traditional methods to decompose the total effect into direct and indirect effects involve comparing regression coefficients before and after controlling for the potential mediator; however, these methods are inappropriate for use with non-linear models^2^. The Buis method decomposes the total causal effect of a categorical variable within a logistic regression into indirect and direct effects, and it does not require a normally distributed mediator variable. It has also been applied to the Virus Watch cohort to decompose the total effect of occupation on SARS-CoV-2 serological status^3^.

The direct effect was the association between migration status and SARS-CoV-2 infection due to all other causes not accounted for in the model. It was derived by comparing the proportion of migrants with evidence of infection with the counterfactual proportion of UK-born individuals who would have had evidence of infection if they had the same distribution of household overcrowding status as the migrant individuals i.e., household overcrowding status was kept constant. To estimate the direct effect, confounders of both the exposure-mediator, mediator-outcome and exposure-outcome mediator must be adjusted for. Based on the DAG in Supplementary Figure 1, the minimally sufficient adjustment set for the direct effect comprises age at baseline, sex at birth, ethnicity, clinical vulnerability, total household income at baseline and occupation.

The indirect effect was the association between migration status and SARS-CoV-2 infection due to differences in household overcrowding status. It is derived by comparing the proportion of migrants who had evidence of SARS-CoV-2 infection with the counterfactual proportion of migrants who would have had evidence of infection if they had the same distribution of household overcrowding status as UK-born participants, while adjusting for the same confounders that were adjusted for when estimating the direct effect. The total effect is estimated by summing the indirect and direct coefficients on the log scale.

Model checks for multicollinearity, particularly in the case of migration status and ethnicity, were conducted using logistic regression controlled for age, sex and ethnicity with the *glm* command in R version 4.1.2 with the family set to binomial and a logit link. The generalised variance inflation factor (GVIF) was calculated using the *vif* command in the *car* package^4^ with adjustment for the number of coefficients in the variables^5^. GVIFs adjusted for the number of coefficients in the variables were all below 10 (Table 2 below), which suggests a lack of evidence of multicollinearity^6^.

**Table 2: GVIFs adjusted for the number of coefficients in the variable for a regression model representing the total effect**

| **Variable** | **GVIF adjusted for number of coefficients in the variable** |
| --- | --- |
| UK born status | 1.40 |
| Age group | 1.01 |
| Sex | 1.14 |
| Ethnicity | 1.12 |

### Supplementary Box 3: Variables used for multiple imputation

The following variables were included as predictor variables:

- Index of multiple deprivation
- Household region
- Sex
- Age at study entry
- Ethnicity
- UK-born status
- Whether there are children living in the household
- Accommodation-related variables: type, self-contained status, presence of central heating, ownership/rental status, number of rooms in household and presence of damp or mould
- Household overcrowding status
- Combined household income
- Clinical vulnerability status
- Living with children
- Occupation
- Whether an individual had a positive test in the study period

### Supplementary Table 1: Self-reported ethnicity of individuals with a missing country of birth

| **Ethnicity** | **n (%)** |
| --- | --- |
| White British | 1,986 (73) |
| White Irish | 57 (2.1) |
| White Other | 116 (4.2) |
| Mixed | 34 (1.2) |
| South Asian | 58 (2.1) |
| Other Asian | 21 (0.8) |
| Black | 13 (0.5) |
| Other | 7 (0.3) |
| Missing | 438 (16) |

### Supplementary Table 2: STROBE checklist (cohort study specific)

|  | Item No | | Recommendation | Page No |
| --- | --- | --- | --- | --- |
| **Title and abstract** | 1 | | (*a*) Indicate the study’s design with a commonly used term in the title or the abstract | p1, 2 |
|  |  |  | (*b*) Provide in the abstract an informative and balanced summary of what was done and what was found |  |
| Introduction | | | | |
| Background/rationale | 2 | | Explain the scientific background and rationale for the investigation being reported | p4 |
| Objectives | 3 | | State specific objectives, including any prespecified hypotheses | p4 |
| Methods | | | | |
| Study design | 4 | | Present key elements of study design early in the paper | p5 |
| Setting | 5 | | Describe the setting, locations, and relevant dates, including periods of recruitment, exposure, follow-up, and data collection | p5 |
| Participants | 6 | | (*a*) Give the eligibility criteria, and the sources and methods of selection of participants. Describe methods of follow-up | p5 |
|  |  |  | (*b*) For matched studies, give matching criteria and number of exposed and unexposed |  |
| Variables | 7 | | Clearly define all outcomes, exposures, predictors, potential confounders, and effect modifiers. Give diagnostic criteria, if applicable | p5,6 |
| Data sources/ measurement | 8* | | For each variable of interest, give sources of data and details of methods of assessment (measurement). Describe comparability of assessment methods if there is more than one group | p5,6 |
| Bias | 9 | | Describe any efforts to address potential sources of bias | p7 |
| Study size | 10 | | Explain how the study size was arrived at | p5 |
| Quantitative variables | 11 | | Explain how quantitative variables were handled in the analyses. If applicable, describe which groupings were chosen and why | p5,6 |
| Statistical methods | 12 | | (*a*) Describe all statistical methods, including those used to control for confounding | p6,7 |
|  |  |  | (*b*) Describe any methods used to examine subgroups and interactions |  |
|  |  |  | (*c*) Explain how missing data were addressed |  |
|  |  |  | (*d*) If applicable, explain how loss to follow-up was addressed |  |
|  |  |  | (*e*) Describe any sensitivity analyses |  |
| Results | | | |  |
| Participants | 13* | | (a) Report numbers of individuals at each stage of study—eg numbers potentially eligible, examined for eligibility, confirmed eligible, included in the study, completing follow-up, and analysed | Figure 1, p8 |
|  |  |  | (b) Give reasons for non-participation at each stage |  |
|  |  |  | (c) Consider use of a flow diagram |  |
| Descriptive data | 14* | | (a) Give characteristics of study participants (eg demographic, clinical, social) and information on exposures and potential confounders | Table 1, p10-16 |
|  |  |  | (b) Indicate number of participants with missing data for each variable of interest |  |
|  |  |  | (c) Summarise follow-up time (eg, average and total amount) |  |
| Outcome data | 15* | | Report numbers of outcome events or summary measures over time | Table 2, p17  Table 3, p18-21 |
| Main results | 16 | (*a*) Give unadjusted estimates and, if applicable, confounder-adjusted estimates and their precision (eg, 95% confidence interval). Make clear which confounders were adjusted for and why they were included | | Table 4, p22 |
|  |  | (*b*) Report category boundaries when continuous variables were categorized | |  |
|  |  | (*c*) If relevant, consider translating estimates of relative risk into absolute risk for a meaningful time period | |  |
| Other analyses | 17 | Report other analyses done—eg analyses of subgroups and interactions, and sensitivity analyses | | p22,23 |
| Discussion | | | | |
| Key results | 18 | Summarise key results with reference to study objectives | | p24 |
| Limitations | 19 | Discuss limitations of the study, taking into account sources of potential bias or imprecision. Discuss both direction and magnitude of any potential bias | | p24, 25 |
| Interpretation | 20 | Give a cautious overall interpretation of results considering objectives, limitations, multiplicity of analyses, results from similar studies, and other relevant evidence | | p24, 25 |
| Generalisability | 21 | Discuss the generalisability (external validity) of the study results | | p24, 25 |
| Other information | | | | |
| Funding | 22 | Give the source of funding and the role of the funders for the present study and, if applicable, for the original study on which the present article is based | | p27 |

*Give information separately for exposed and unexposed groups.

### Supplementary Figure 1: Directed acyclic graph of the relationship between migration status and SARS-CoV-2 infection

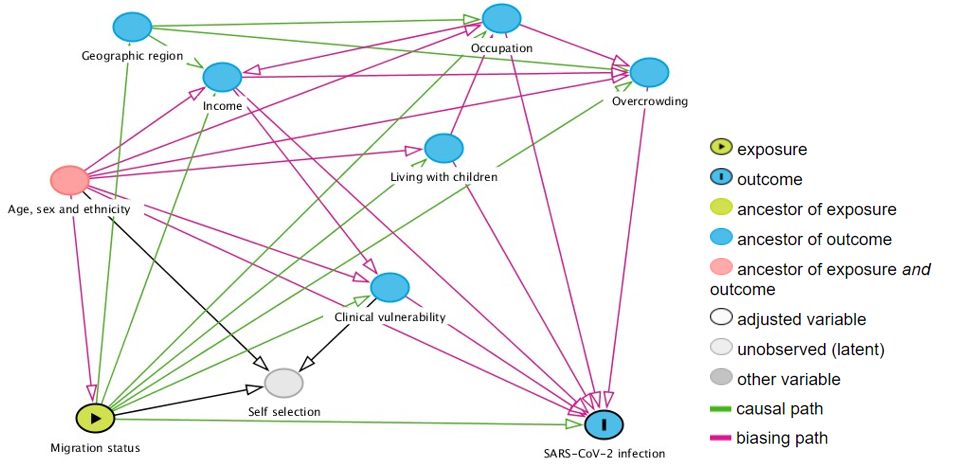
